# Interplay Of Serum Bilirubin and Tobacco Smoking with Lung and Head and Neck Cancers in a Diverse, EHR-linked Los Angeles Biobank

**DOI:** 10.1101/2023.09.29.23296364

**Authors:** Vidhya Venkateswaran, Ella Petter, Kristin Boulier, Yi Ding, Arjun Bhattacharya, Bogdan Pasaniuc

**Affiliations:** Department of Pathology and Laboratory Medicine, David Geffen School of Medicine, University of California, Los Angeles, Los Angeles, CA 90095, USA; Department of Oral Biology, School of Dentistry, University of California, Los Angeles, Los Angeles, CA 90095, USA; Bioinformatics Interdepartmental Program, University of California, Los Angeles, Los Angeles, CA 90095, USA; Department of Human Genetics, David Geffen School of Medicine, University of California, Los Angeles, Los Angeles, CA 90095, USA; Department of Computational Medicine, David Geffen School of Medicine, University of California, Los Angeles, Los Angeles, CA 90095, USA; Institute for Precision Health, University of California, Los Angeles, Los Angeles, CA 90095, USA; Department of Medicine, Division of Cardiology, University of California, Los Angeles, Los Angeles, CA 90095, USA; Department of Computer Science, University of California, Los Angeles, Los Angeles, CA 90095, USA; Department of Epidemiology, University of Texas MD Anderson Cancer Center, Houston, TX 77030, USA

**Author notes:** Corresponding author Vidhya Venkateswaran, Department of Pathology & Laboratory Medicine Geffen School of Medicine at UCLA, 10833 Le Conte Ave, CHS 33-365 Los Angeles, CA 90024. Equal contribution (Arjun Bhattacharya, Bogdan Pasaniuc). Conflict of interest statement: The authors declare no potential conflicts of interest.

## Abstract

**Background:** Bilirubin is a potent antioxidant with a protective role in many diseases. We examined the relationships between serum bilirubin (SB) levels, tobacco smoking (a known cause of low SB), and aerodigestive cancers, grouped as lung (LC) and head and neck (HNC).

**Methods:** We examined the associations between SB, LC and HNC using data from 393,210 participants from UCLA Health, employing regression models, propensity score matching, and polygenic scores.

**Results:** Current tobacco smokers showed lower SB (-0.04mg/dL, 95% CI: [-0.04, -0.03]), compared to never-smokers. Lower SB levels were observed in HNC and LC cases (-0.10 mg/dL, [-0.13, -0.09] and -0.09 mg/dL, CI [-0.1, -0.07] respectively) compared to cancer-free controls with the effect persisting after adjusting for smoking. SB levels were inversely associated with HNC and LC risk (ORs per SD change in SB: 0.64, CI [0.59,0.69] and 0.57, CI [0.43,0.75], respectively). Lastly, a polygenic score (PGS) for SB was associated with LC (OR per SD change of SB-PGS: 0.71, CI [0.67, 0.76]).

**Conclusions:** Low SB levels are associated with an increased risk of both HNC and LC, independent of the effect of tobacco smoking with tobacco smoking demonstrating a strong interaction with SB on LC risk. Additionally, genetically predicted low SB (from polygenic scores) is negatively associated with LC.

**Impact:** These findings suggest that SB could serve as a potential early biomarker for LC and HNC.

## Introduction

Bilirubin is a compound found in the blood, derived from the breakdown of heme, a component of hemoglobin found in red blood cells. At the end of their life cycle, red blood cells are broken down in the spleen and bone marrow, forming bilirubin, which is then transported to the liver and excreted in bile. Thus, the level of serum bilirubin (SB) is an indicator of liver health with high SB levels used as markers of liver disease^1, 2^. However, low levels of SB are not currently recognized to have clinical significance and do not factor into diagnosis or treatment planning in a majority of diseases^1^. Recent studies, however, have redefined the role of SB in the human body, recognizing it as a metabolic hormone with potent antioxidant effects^3^. Low SB levels are associated with increased risk of diseases including cancers, metabolic and cardiovascular diseases^4–6^. Additionally, low SB levels have been associated with poorer survival and worse prognosis in lung and oral cancers although these thresholds vary between studies and the type of bilirubin measured ^7, 8^.

Tobacco smoking is a commonly reported factor contributing to reduced SB levels, attributed to the oxidative stress induced by tobacco smoke, which disrupts the typical bilirubin metabolism within the body^9^. Tobacco smoking is also a well-established risk factor for aerodigestive cancers, i.e. lung cancer (LC) and head and neck cancers (HNC)^10^. Studies have reported inverse associations between SB and LC with these associations persisting after accounting for tobacco use^11, 12^. Studies have also reported interactions between SB and tobacco smoking on the risk of LC^11, 12^. However, the associations between SB and HNC remain understudied.

In this study, we examined the interplay of SB and tobacco smoking with aerodigestive cancers - HNC and LC, using lab values and electronic health records (EHR) information from a hospital-based biobank in Los Angeles (***Supplementary*** Figure 1). We found that SB levels were inversely associated with tobacco smoking in this study population. We also observed inverse associations between SB levels and both HNC and LC with these associations persisting after adjusting for tobacco use. SB significantly interacted with tobacco on the risk of LC, but not HNC. Lastly, we found that a polygenic score for SB is inversely correlated with LC.

## Materials and Methods

### Study Population

The study population is derived from the UCLA Discovery Data Repository (DDR) which consists of de-identified patient electronic health records (EHR) of UCLA patients, including data on demographics, diagnostic codes, laboratory values, prescription data, and procedure codes^13^. The broad inclusion criteria in this study were 1) participants aged 18 and above, 2) participants with at least one result for a total SB laboratory test . HNC and LC cases (ascertainment detailed below under ‘HNC, LC, and Tobacco Smoking Ascertainment’) were identified through ICD-codes in the EHR and controls were participants without any ‘malignancy’ or ‘cancer’ diagnoses in their EHR. 393,210 participants met the inclusion criteria and were included in the analyses. (***Supplementary Figure 2***)

For the polygenic score analysis, we included participants whose genotypes were available in the UCLA ATLAS biobank^14, 15^. These participants were genotyped on a custom array^16^ and underwent stringent genotype quality control including removal of strand ambiguous SNPs and variants with >5% missingness and filtering out SNPs that did not pass the Hardy-Weinberg equilibrium test with a p-value set at “--hwe 0.001”. The QC-ed genotypes were imputed to the TOPMed Freeze5 referencer^17, 18^. Variants at the threshold of R2 > 0.90 and minor allele frequency > 1% were filtered out from analyses. All quality control steps were conducted using PLINK 1.925^19^. We computed the top 10 principal components for the study population using FlashPCA2 software^20^ and grouped the genotyped study population into genetically inferred ancestry groups (GIAs) - European American (EA), Hispanic/Latin American (HL), East Asian American (EAA), African American (AA) - by k-nearest neighbor (KNN) stratification of the principal components, using the continental ancestry populations from 1000 Genomes Project as a reference^21, 22^.

For the polygenic score analysis, we restricted the analyses to participants of European American GIA to match the training population of the publicly available polygenic score used in this study (detailed below under ‘Polygenic score selection and calculation’). 15,023 participants met the inclusion for the polygenic score analysis criteria and were included in this analysis.

### Bilirubin Biomarker Measurements

The primary biomarker analyzed in this study is serum total bilirubin (SB), measured in mg/dL. The SB measurements were extracted from the patient’s laboratory results within the deidentified EHR database (DDR). Since most patients have multiple results over their encounter history for SB laboratory tests, we computed the mean of each patient’s maximum and minimum total bilirubin values to get a mid-range total SB value for each individual. To filter out errors in data entry and significant outliers, we excluded participants with SB values that were less than the 0.001th percentile or greater than the 95th percentile. This step also served to exclude patients with consistently high bilirubin values secondary to liver disease or medication side effects. We further excluded participants whose SB was recorded as ‘999999’ which corresponds to NA values. (***Supplementary*** Figure 2)

### HNC, LC, and Tobacco Smoking Ascertainment

Smoking history was extracted from the patient’s self-reported information. We binarized participants to ever- and never smokers based on their smoking history.

- Ever-smokers included participants who selected any of the following options in their health history: ‘Former’, ‘Heavy Smoker’, ‘Light Smoker’, ‘Smoker, Current Status Unknown’, and ‘Some Days’.
- Never smokers included participants who selected the following options: ‘Never’, ‘Passive Smoke Exposure - Never Smoker’.

We excluded participants who were assigned ‘Never Assessed’ or whose smoking history was recorded as ‘Unknown’. Further, we created a ‘Current Smoker’ label for participants who answered selected ’Every Day’, ‘Some Days’, ‘Light Smoker’, or ‘Heavy Smoker’ in their smoking history question. This variable was used for additional analysis of the association between tobacco smoking and SB levels. (***Supplementary*** Figure 2)

Head and neck cancer (HNC) and lung cancer (LC) status were ascertained by extracting ICD codes from the participant’s EHR. A detailed list of the ICD codes used to ascertain HNC and LC status is included in the ***Supplementary Table 1***. Participants were designated as a ‘HNC Case’ or ‘LC Case’ if the ICD codes for HNC or LC were present in their EHR respectively. Participants were assigned to ‘Controls’ when their EHR did not contain any ICD codes for ‘malignancy’ or ‘cancers’.

### Polygenic score selection and calculation

A publicly available polygenic score (PGS) for ‘Total Bilirubin’ was used for polygenic score computation (PGS002160 from the PGS Catalog^23, 24^). This PGS was trained using 391,124 European individuals from the UK biobank and includes 120,068 SNPs. PGS training analyses were adjusted for sex, age, birth date, Townsend’s deprivation index, and the first 16 principal components of the genotype matrix to account for population stratification. We computed the PGS for each genotyped UCLA ATLAS biobank participant by multiplying the individual risk allele dosages by their corresponding weights provided in the PGS catalog using the ‘pgs_calc’ pipeline^24^. The PGS was mean-centered and standardized by the standard deviation within the European American genetic ancestry group to generate a PGS Z-score which was used in further analysis (n = 15,023). We restricted the PGS analysis to European American ancestry group since the original PGS was trained in Europeans, and studies have shown that the predictive performance of PGS is unreliable when used in ancestries that are genetically dissimilar to the trained population^25^.

### Statistical Analyses

All analyses were conducted in either Python 2.6.8^26^ or R 4.2.1^27^. We used linear regression models to evaluate the associations between the dependent variable of SB levels and independent variables of tobacco smoking, HNC, and LC. We adjusted for participant age, sex, and self-reported race/ethnicity (SIRE) in all models. In a subsequent model to evaluate cancer effect sizes on SB, we additionally accounted for tobacco smoking by including it as an independent variable. Linear coefficients and confidence intervals were calculated, with P-values from Wald-type test statistics.

Propensity scores were estimated by logistic regression analysis, with cancer status as the dependent variable and age, sex, SIRE, and tobacco smoking as independent variables. We used 1:1 propensity matching to create more balanced analysis groups, creating two sets of groups: 2,037 HNC cases and 2,037 HNC controls, and 2,373 LC cases and 2,373 LC controls with no significant differences in age, sex, SIRE or tobacco smoking.

To evaluate the effect of observed SB and genetically predicted SB on HNC and LC, we used logistic regression models with HNC or LC as the dependent variable and SB as the independent variable. The SB variable was the observed SB levels from the EHR for the observed SB analysis and the computed polygenic score for SB for the genetically predicted SB analysis. These models were adjusted for age, sex, SIRE and tobacco smoking. Odds ratios and confidence intervals were calculated, with P-values from Wald-type test statistics for these analyses.

### Ethical Approval

Patient Recruitment and Sample Collection for Precision Health Activities at UCLA is an approved study by the UCLA Institutional Review Board (UCLA IRB #17-001013). All participants provided informed consent to participate in the research.

## Results

### Effect of demographic factors and tobacco smoking on SB

We used a multivariable linear regression model to systematically evaluate the differences in SB level distributions among demographic factors and tobacco smoking. We find that males on average had 0.15 mg/dL higher SB compared to females *(CI [0.146, 0.151])*, and older individuals had a 0.0009 mg/dL increase in SB, per year of age, CI [0.0009,0.001]). When compared to individuals who self-reported as ‘White’, individuals who self-reported as ‘Asian’ had on average 0.02 mg/dL higher SB values, *(CI [0.015, 0.023])* and as ‘African American’ had 0.01 mg/dL lower SB values *(CI [-0.02, -0.005])*. Lastly, individuals who self-identified as ‘Hispanic/Latin American’ ethnicity had 0.02 mg/dL higher SB compared to individuals who self-identified as ‘Not Hispanic/Latin American’ *CI [0.02, 0.03]* (***Table 1***).

Next, we evaluated the association between SB levels and tobacco smoking. In a linear regression model adjusting for age, sex, and SIRE, ever-smokers demonstrated an inverse association with SB (*-0.017 mg/dL, CI [-0.019, -0.014]*). This effect size persists and increases when comparing ‘current smokers’ (*-0.038 mg/dL, CI [-0.043, -0.032]*) to ‘never smokers’ ***(Table 2)*.**

These results demonstrate that SB levels vary by sex, age and SIRE. Additionally, tobacco smoking demonstrates an inverse relationship with SB levels with a larger effect size noted in current smokers. These results replicate the results of other studies that have reported the inverse association between SB and tobacco smoking.

The baseline characteristics of 393,210 participants with 2,039 HNC cases, 2,378 LC cases, and 388,793 cancer controls are shown in ***Supp Table 2.***

**Table 1:** Effects of demographic factors on SB. Results of a multivariable linear regression model adjusted for age, sex, and SIRE to evaluate the associations between these factors and SB levels which was the dependent variable. P value significance threshold was set at 0.05/8 to account for multiple testing.

| Reference | Variables | Effect Size | 95% CI | P-Value |
| --- | --- | --- | --- | --- |
| Female | Male | 0.15 | 0.146,0.151 | < 0.0001 |
|  | Patient Age | 0.001 | 0.0009,0.001 | < 0.0001 |
| 'White' | 'American Indian or<br>Alaska Native' | -0.02 | -0.03,-0.002 | 0.03 |
| 'White' | 'Asian' | 0.02 | 0.015,0.02 | < 0.0001 |
| 'White' | 'Black or African<br>American' | -0.01 | -0.016,-0.004 | 0.0004 |
| 'White' | 'Native Hawaiian or<br>Other Pacific<br>Islander' | 0.03 | -0.004,0.06 | 0.09 |
| 'White' | 'Other Race' | 0.004 | -0.0001,0.009 | 0.06 |
| 'Not Hispanic or<br>Latin ethnicity' | 'Hispanic or Latin<br>ethnicity' | 0.02 | 0.019,0.03 | < 0.0001 |

**Table 2:** Effect of Tobacco Smoking on SB. Results of linear regressions adjusted for age, sex, SIRE to evaluate the associations between tobacco smoking and SB levels (dependent variable).

| Reference | Variables | Effect on SB<br>mg/dL | 95% CI | P-Value |
| --- | --- | --- | --- | --- |
| Never smokers | Ever-smokers | -0.017 | -0.019, -0.014 | < 0.0001 |
| Never smokers | Current smokers | -0.038 | -0.043, -0.032 | < 0.0001 |

### Low SB is associated with HNC and LC independent of tobacco smoking

Given the inverse association between tobacco smoking and SB levels, we next evaluated the association between SB and HNC/LC. First, we examined the association between SB and HNC/LC, with LC or HNC case-control status as the dependent variable. In two separate logistic regression models, SB demonstrates a protective association on both HNC and LC, after adjusting for age, sex, SIRE, and tobacco smoking *(ORs: 0.64, CI [0.59, 0.69] and 0.71, CI [0.67, 0.76]) respectively per SD of SB)*. Additionally, interaction terms of SB and tobacco smoking demonstrated significant effects on LC but not on HNC *(OR per SD: 0.79, CI [0.70, 0.89]) and 0.98, CI [0.85,1.14] respectively). (****Table 3*).**

Next, we evaluated the associations between SB and HNC and SB and LC independently in separate generalized linear models with SB as the dependent variable (i.e. opposite direction of the previous analysis), to obtain effect sizes of the cancers on SB levels. HNC and LC cases both independently demonstrate inverse associations with SB, after adjusting for age, sex, and SIRE (*-0.11 mg/dL, CI [-0.13, -0.1] and -0.09 mg/dL, CI [-0.1, -0.07], respectively*) when compared to cancer-free controls. These inverse associations between HNC/LC, and SB persist after adjusting for smoking history (*-0.11 mg/dL, CI [-0.13, -0.09] and -0.08 mg/dL, CI [-0.1, -0.07], respectively*). (***Supplementary Table 3***).

Lastly, we used propensity score matching for HNC and LC to create balanced comparison groups and to evaluate the association between SB and risk of HNC/LC. The baseline characteristics of the propensity-matched HNC and LC groups are shown in ***Supplementary Tables 4a and 4b***. The two sets of groups were matched on age, sex, SIRE and tobacco smoking, differing only in HNC status or LC status. In two separate linear regression models, HNC and LC individually demonstrated inverse associations with SB *(-0.11mg/dL, CI [-0.13, -0.09]) and -0.083mg/dL, CI [-0.10, -0.07] respectively)* in this analysis. These results replicate the results from the linear regression model above.

Overall, SB demonstrates inverse associations with both HNC and LC that are likely mediated by factors other than tobacco smoking. Tobacco smoking also has a significant interaction with SB on lung cancer risk, suggesting that the inverse association between LC and SB is exacerbated by smoking with almost double the effect size of SB alone on LC. A similar trend is not observed for HNC.

**Table 3:** Effects of SB levels on HNC and LC. Results of logistic regressions adjusted for age, sex, SIRE and tobacco smoking to evaluate the associations between HNC/LC (dependent variables) and SB levels.

|  | OR per SD of SB | 95% CI | P-Value |
| --- | --- | --- | --- |
| **Outcome - HNC** |  |  |  |
| SB | 0.64 | 0.59,0.69 | < 0.0001 |
| SB and smoking interaction variable | 0.98 | 0.85, 1.14 | 0.8 |
| **Outcome - LC** |  |  |  |
| SB | 0.71 | 0.67, 0.76 | < 0.0001 |
| SB and smoking interaction variable | 0.79 | 0.70, 0.89 | 0.0002 |

### Polygenic score for SB (SB-PGS) is correlated with LC

Having established the inverse association between SB and both LC and HNC, we next used a publicly-available polygenic score for SB to assess if a genetic predisposition to elevated SB levels correlated with LC and HNC risk. In a subgroup of genotyped European GIA patients from the UCLA ATLAS biobank (baseline characteristics available in ***Supplementary Table 5***), first, we validated a PGS for total serum bilirubin (SB-PGS) using observed SB values from the EHR. The SB-PGS demonstrated a strong association with observed SB levels (*0.094 mg/dL per standard deviation increase in SB-PGS, CI [0.091, 0.097*], demonstrating reliability in predicting SB within the European American ancestry subgroup. In this subgroup, after adjusting for age, sex, smoking history and the first 5 principal components, SB-PGS demonstrated inverse associations with LC (*n=124 cases, OR: 0.78, CI [0.65,0.94]*) but not HNC, *(n = 152 cases; OR = 1.01, CI [0.86,1.19])*. These results suggest that the genetic control of SB could potentially play a role in lung cancer risk.

## Discussion

In this study, we analyzed the associations between SB, tobacco smoking and aerodigestive cancers - HNC and LC, using data from the UCLA DDR and EHR-linked biobank. We found that SB was inversely associated with HNC and LC, independent of tobacco smoking. Tobacco smoking significantly interacted with SB on LC risk. Lastly, observed SB was associated with both HNC and LC while genetically predicted SB was only associated with LC.

This is the first large-scale study to report that low SB is associated with HNC. The inverse association between SB and LC noted in our study replicates the results of other studies on SB and LC^7–11^. A potential mechanism for this inverse association is likely through the antioxidant and anti-inflammatory action of SB on known cancer-promoters such as reactive oxygen species and inflammatory factors^25--27^. We evaluated two factors that could affect the associations between SB and these two cancers: tobacco smoking and the genetically-predicted SB levels.

Tobacco smoking is a well-known contributor of free radicals and reactive oxygen species, playing an important role in carcinogenesis^28^. Tobacco smoking demonstrated a negative association with SB in this study with persistent SB-HNC/LC associations observed after adjusting for tobacco smoking. These results suggest that the associations between SB-HNC/LC are not solely driven by tobacco smoking. In fact, tobacco smoking interacted with SB to increase LC risk with the effect size almost equal to that of SB alone. The difference in the SB-tobacco smoking interaction results between HNC and LC could likely be explained by differences in etiology (alcohol consumption and human papillomavirus infection for HNC), differential tissue response, local environment, and microbiome differences between the two sites. From a clinical perspective, these results suggest that SB could be an early, low-cost laboratory biomarker for HNC and LC in combination with other risk factors even as further studies are needed to understand these potential pathways and to validate these findings. We should also note that current laboratory tests for SB are not precise enough to capture small differences and further work is needed to more precisely measure and threshold SB.

Next, an evaluation of the genetic control of SB and its effect on these two cancers revealed an inverse association with LC, suggesting the possible influence of genetic control of SB on the susceptibility to LC. In fact, a recent study used Mendelian Randomization methods to evaluate the association between two genetic variants which account for ∼40% of population-level SB variability and LC and found evidence of a causal association^29^ between SB and LC. The inability of the SB-PGS to predict HNC could be attributed to the multifactorial nature of HNC development or that the variants in the SB-PGS have very small effects on HNC that we could not capture. Further validation studies are required with adequate sample sizes across the subtypes of HNC and including information on a larger spectrum of environmental risk factors, including alcohol consumption and human papillomavirus infection status. Although multiple stages of research must be completed before clinical translation is possible and equitable; such as ensuring the robust predictive performance of SB-PGS across diverse ancestral groups, polygenic scores for SB may hold promise in risk-stratifying LC in combination with other scores.

The main strengths of our study are that we used data from an hospital based EHR and biobank, thus including patients from a real-world hospital system of diverse racial and ethnic groups. The large sample size adequately powered the analysis of SB, HNC, and LC. Given the significant role that tobacco smoking plays in the development of HNC and LC, we thoroughly evaluated the effects of tobacco smoking on SB levels and potential interactions with SB levels in the context of the cancer risk. Lastly, we used a polygenic score to explore the role that genetics may play in SB levels and HNC/LC risk, to identify potential genetic biomarkers that could be used to predict cancer.

We end this discussion with some limitations of our study to be considered when interpreting and applying the results of this study. The observational nature of our study prevents us from making causal interpretations and assigning directions to the associations observed between SB levels, tobacco smoking, HNC/LC. Next, the de-identified nature of the data source prevented us from including socioeconomic variables that could have effects on tobacco smoking and cancer risk. The population included in a hospital-based data source might demonstrate fundamental differences from other individuals, such as better healthcare access. Lastly, we lacked information on cancer subtype, staging, and grading and could not investigate these factors that could yield more information on the potential effects of low SB through the course of the cancer.

## Data Availability

All data produced in the present work are contained in the manuscript

https://github.com/drvidhya/bili_hnc_lc

## Acknowledgments

VV is funded by NIH/NIDCR 5K12DE027830. The ATLAS Community Health Initiative is supported by UCLA Health, the David Geffen School of Medicine, and a grant from the UCLA Clinical and Translational Science Institute (UL1TR001881). KB is funded by an NIH 5T32H007895 grant. BP was partially supported by NIH awards R01 HG009120, R01 MH115676, R01 CA251555, R01 AI153827, R01 HG006399, R01 CA244670, and U01 HG011715.

We gratefully acknowledge the resources provided by the Institute for Precision Health (IPH) and participating UCLA ATLAS Community Health Initiative patients. The UCLA ATLAS Community Health Initiative and UCLA ATLAS Precision Health Biobank, is a program of the IPH, which directs and supports the biobanking and genotyping of biospecimen samples from participating UCLA patients in collaboration with the David Geffen School of Medicine, UCLA Clinical and Translational Science Institute grant number UL1TR001881, and UCLA Health.

## Data Sharing

All shareable data produced in the present work are contained in the manuscript. The code used to generate the results are available at https://github.com/drvidhya/bili_hnc_lc.

## Notes

### Competing Interest Statement

The authors have declared no competing interest.

