## Supplemental Figures and Tables for "Interplay Of Serum Bilirubin and Tobacco Smoking with Lung and Head and Neck Cancers in a Diverse, EHR-linked Los Angeles Biobank"

### Supplementary Tables and Figures

**Supplementary Figure 1: Directed acyclic graph representing the associations between tobacco smoking and serum total bilirubin with dotted lines representing the associations evaluated in this study**

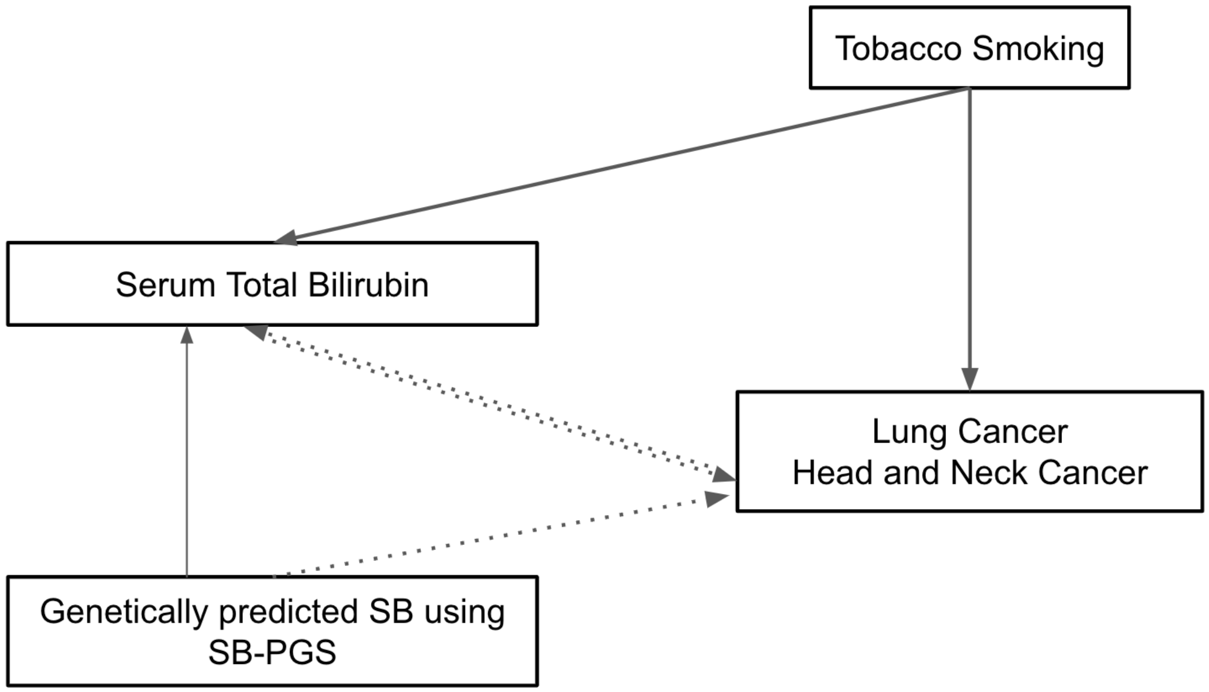

**Supplementary Figure 2: Flowchart of participant inclusion and exclusion**

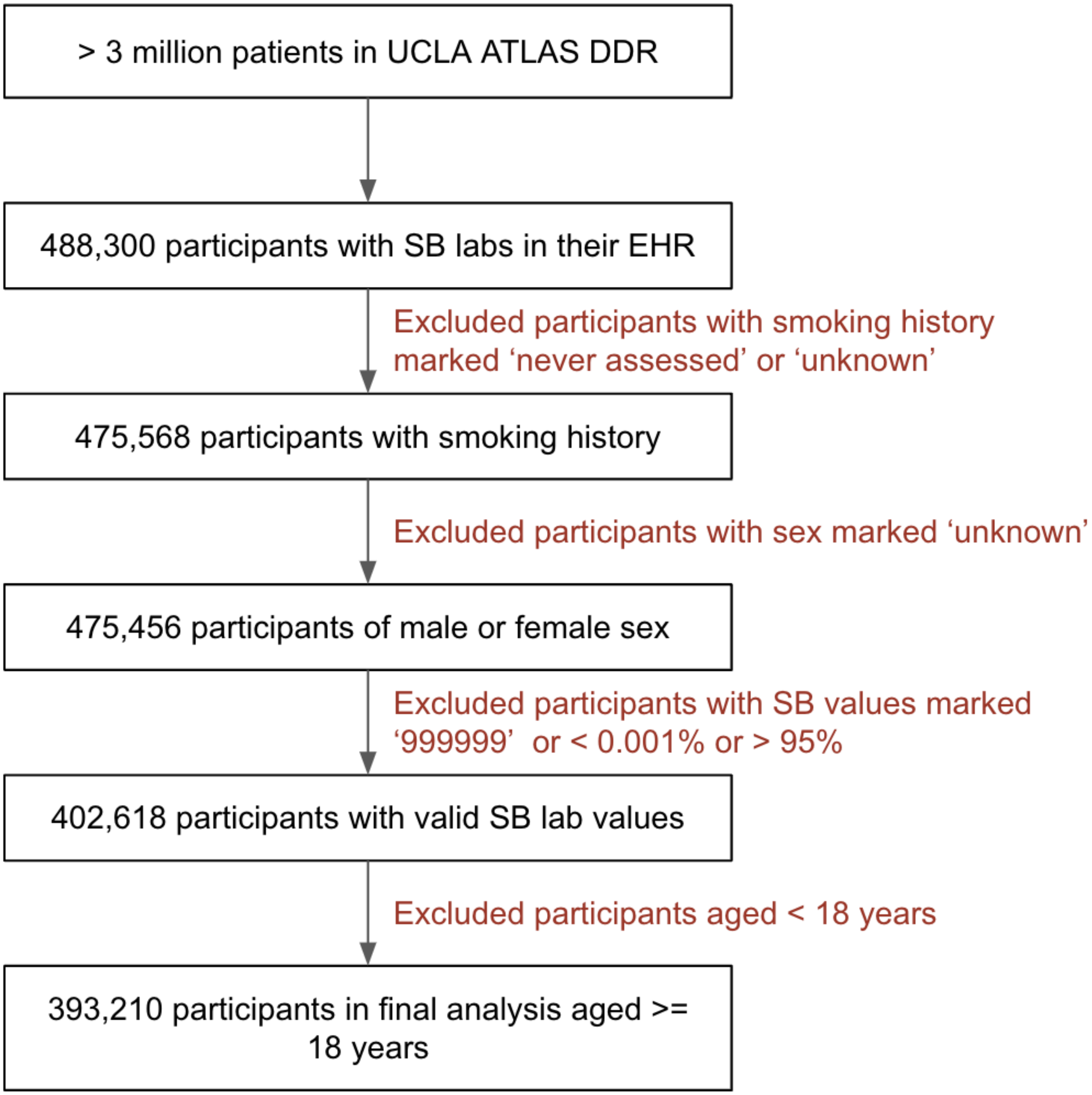

**Supplementary Table 1: ICD Codes for Head and Neck Cancer and Lung Cancer**

| Type of Cancer | ICD Codes |
|---|---|
| Head and Neck Cancer | '140', '140.1', '140.3', '140.4', '140.5', '140.6', '140.8', '140.9', '141', '141.1', '141.2', '141.3', '141.4', '141.5', '141.6', '141.8', '141.9', '142', '142.1', '142.2', '142.8', '142.9','143', '143.1', '143.8', '143.9', '144', '144.1', '144.8', '144.9', '145', '145.1', '145.2', '145.3', '145.4', '145.5', '145.6', '145.8', '145.9', '146', '146.1', '146.2', '146.3', '146.4', '146.5', '146.6', '146.7', '146.8', '146.9', '147', '147.1', '147.2', '147.3', '147.8', '147.9’, '148', '148.1', '148.2', '148.3', '148.8', '148.9', '149', '149.1', '149.8', '149.9', '160', '160.1', '160.2', '160.3', '160.4', '160.5', '160.8', '160.9’, '161', '161.1', '161.2', '161.3', '161.8', '161.9', 'C00.0', 'C00.1', 'C00.2', 'C00.3', 'C00.4', 'C00.5', 'C00.6', 'C00.8', 'C00.9', 'C02.0', 'C02.1', 'C02.2', 'C02.3', 'C02.4', 'C02.8', 'C02.9', 'C06.0', 'C06.1', 'C06.2', 'C06.8', 'C06.80', 'C06.89’, 'C06.9', 'C08.0', 'C08.1', 'C08.9', 'C09.0', 'C09.1', 'C09.8', 'C09.9', 'C10.0', 'C10.1', 'C10.2', 'C10.3', 'C10.4', 'C10.8', 'C10.9', 'C11.0', 'C11.1',”C11.2', 'C11.3', 'C11.8', 'C11.9', 'C13.0', 'C13.1', 'C13.2', 'C13.8', 'C13.9', 'C14.0','C14.2', 'C14.8', 'C30.0'. |

| | |
|---|---|
| Lung Cancer | 'C34.0', 'C34.1', 'C34.2', 'C34.3', 'C34.8', 'C34.9','162.0', '162.2', '162.3', '162.4', '162.5', '162.8', '162.9' |

**Supplementary Table 2: Baseline characteristics of participants, stratified by case: control status**

| | | Cancer Controls | LC Cases | HNC Cases |
|---|---|---|---|---|
| n | | 388793 | 2378 | 2039 |
| Patient Age, mean (SD) | | 57.0 (18.7) | 75.8 (10.9) | 69.7 (13.5) |
| SB mg/dL, mean (SD) | | 0.6 (0.3) | 0.5 (0.2) | 0.5 (0.2) |
| Sex, n (%) | Female | 213979 (55.0) | 1259 (52.9) | 589 (28.9) |
| | Male | 174814 (45.0) | 1119 (47.1) | 1450 (71.1) |
| Smoking History, n (%) | Never Smokers | 270277 (69.6) | 831 (34.9) | 1010 (49.5) |
| | Ever Smokers | 118516 (30.4) | 1547 (65.1) | 1029 (50.5) |
| Self-reported Race, n (%) | American Indian or Alaska Native | 2045 (0.7) | 10 (0.5) | 6 (0.4) |
| | Asian | 36632 (13.4) | 395 (19.6) | 208 (12.8) |
| | Black or African American | 15860 (5.8) | 111 (5.5) | 59 (3.6) |
| | Native Hawaiian or Other Pacific Islander | 530 (0.2) | 9 (0.4) | 2 (0.1) |

| | | | | |
|---|---|---|---|---|
| | Other Race | 32185 (11.8) | 203 (10.1) | 171 (10.5) |
| | White or Caucasian | 161596 (59.1) | 1111 (55.2) | 1078 (66.2) |
| Self-Reported Ethnicity, n (%) | Hispanic or Latin | 33631 (14.3) | 136 (7.0) | 129 (9.0) |
| | Not Hispanic or Latin | 211951 (82.2) | 1681 (88.3) | 1380 (87.9) |

**Supplementary Table 3: Effects of HNC and LC on SB**

Results of linear regressions adjusted for age, sex, SIRE to evaluate the associations between HNC/LC and SB levels(dependent variable).

| Variables | Effect on SB mg/dL | 95% CI | P-Value |
|---|---|---|---|
| HNC | -0.11 | -0.13, -0.09 | < 0.0001 |
| HNC (adjusted for smoking) | -0.11 | -0.13, -0.09 | < 0.0001 |
| LC | -0.09 | -0.1, -0.07 | < 0.0001 |
| LC (adjusted for smoking) | -0.08 | -0.1, -0.07 | < 0.0001 |

**Supplementary Table 4a: Propensity matched group baseline characteristics stratified by HNC**

| | | Overall | HNC Controls | HNC Cases |
|---|---|---|---|---|
| n | | 4074 | 2037 | 2037 |
| Patient Age, mean (SD) | | 69.7 (13.5) | 69.7 (13.5) | 69.7 (13.5) |
| Serum Total Bilirubin, [Q1,Q3] | | 0.5 [0.4,0.7] | 0.6 [0.4,0.8] | 0.5 [0.4,0.6] |
| Sex, n (%) | Female | 1174 (28.8) | 587 (28.8) | 587 (28.8) |
| | Male | 2900 (71.2) | 1450 (71.2) | 1450 (71.2) |

| | | | | |
|---|---|---|---|---|
| Smoking History, n (%) | 0 | 2018 (49.5) | 1009 (49.5) | 1009 (49.5) |
| | 1 | 2056 (50.5) | 1028 (50.5) | 1028 (50.5) |
| Self-reported Race, n (%) | American Indian or Alaska Native | 11 (0.3) | 6 (0.4) | 5 (0.3) |
| | Asian | 414 (12.7) | 207 (12.7) | 207 (12.7) |
| | Black or African American | 118 (3.6) | 59 (3.6) | 59 (3.6) |
| | Native Hawaiian or Other Pacific Islander | 4 (0.1) | 2 (0.1) | 2 (0.1) |
| | Other Race | 342 (10.5) | 171 (10.5) | 171 (10.5) |
| | Unknown Race | 210 (6.5) | 105 (6.4) | 105 (6.5) |
| | White or Caucasian | 2156 (66.2) | 1078 (66.2) | 1078 (66.3) |
| Self Reported Ethnicitiy, n (%) | Hispanic or Latin | 343 (9.1) | 173 (9.2) | 170 (9.0) |
| | Not Hispanic or Latin | 3310 (87.9) | 1655 (87.8) | 1655 (88.0) |
| | Unknown Ethnicity | 112 (3.0) | 56 (3.0) | 56 (3.0) |

**Supplementary Table 4b: Propensity matched group baseline characteristics stratified by LC**

| | | Overall | Lung Cancer Controls | Lung Cancer Cases |
|---|---|---|---|---|
| n | | 4746 | 2373 | 2373 |
| Patient Age, mean (SD) | | 75.8 (10.9) | 75.8 (10.9) | 75.8 (10.9) |
| Serum Total Bilirubin, [Q1,Q3] | | 0.5 [0.4,0.7] | 0.5 [0.4,0.7] | 0.5 [0.4,0.7] |
| Sex, n (%) | Female | 2512 (52.9) | 1256 (52.9) | 1256 (52.9) |
| | Male | 2234 (47.1) | 1117 (47.1) | 1117 (47.1) |

| | | | | |
|---|---|---|---|---|
| Smoking History, n (%) | Never Smokers | 1656 (34.9) | 828 (34.9) | 828 (34.9) |
| | Ever Smokers | 3090 (65.1) | 1545 (65.1) | 1545 (65.1) |
| Self Reported Race, n (%) | American Indian or Alaska Native | 15 (0.4) | 5 (0.2) | 10 (0.5) |
| | Asian | 786 (19.6) | 393 (19.6) | 393 (19.6) |
| | Black or African American | 222 (5.5) | 111 (5.5) | 111 (5.5) |
| | Native Hawaiian or Other Pacific Islander | 12 (0.3) | 6 (0.3) | 6 (0.3) |
| | Other Race | 406 (10.1) | 203 (10.1) | 203 (10.1) |
| | Unknown Race | 348 (8.7) | 174 (8.7) | 174 (8.7) |
| | White or Caucasian | 2222 (55.4) | 1111 (55.5) | 1111 (55.3) |
| Self Reported Ethnicitiy, n (%) | Hispanic or Latin | 326 (7.3) | 171 (7.7) | 155 (7.0) |
| | Not Hispanic or Latin | 3918 (88.1) | 1959 (87.8) | 1959 (88.4) |
| | Unknown Ethnicity | 202 (4.5) | 101 (4.5) | 101 (4.6) |

**Supplementary Table 5: Baseline characteristics of European GIA UCLA ATLAS Biobank participants included in SB-PGS analysis**

| | | Cancer Controls | LC Cases | HNC Cases |
|---|---|---|---|---|
| n | | 14871 | 124 | 152 |
| Patient Age, mean (SD) | | 63.2 | 73.8 | 70.1 |
| SB mg/dL, mean (SD) | | 0.6 | 0.5 | 0.5 |
| Sex, n (%) | Female | 7432 (50) | 63 (50) | 30 (20) |

| | | | | |
|---|---|---|---|---|
| | Male | 7439 (50) | 61 (50) | 122 (80) |
| Smoking History, n (%) | Never Smokers | 9421 (64) | 35 (28) | 82 (54) |
| | Ever Smokers | 5450 (36) | 89 (72) | 70 (46) |
